# Zika Virus Infection Knowledge and Communication Preferences Among Women of Reproductive Age in Central Brooklyn, New York: A Thematic Analysis

**DOI:** 10.1101/2024.03.28.24304317

**Authors:** Russell Dowling, Sergios-Orestis Kolokotronis, Azure B. Thompson

**Author notes:** Corresponding author contact information: Russell Dowling. **Author Contributions** Russell Dowling: Conceptualization, data collection, data interpretation, writing. Sergios-Orestis Kolokotronis: Reviewing, editing. Azure Thompson: Supervision, reviewing, editing. **Ethics Statement** This study was reviewed by the SUNY Downstate’s Office of Research Administration Institutional Review Board (IRBNet Number 1617142-2). The study received an exemption under Common Rule Exemption Category 2 [45 CFR 46.104 (d) (i) (ii), or (iii)]: “Research that only includes interactions involving educational tests (cognitive, diagnostic, aptitude, achievement), survey procedures, interview procedures, or observation of public behavior (including visual or auditory recording) if at least one of the following criteria is met: Criterion 3: [45CFR46.104(d)(2)(iii)]: The information obtained is recorded by the investigator in such a manner that the identity of the research participants can readily be ascertained, directly or through identifiers linked to the research participants.”.

## Abstract

The 2016 outbreak of Zika virus (ZIKV) infected millions and resulted in thousands of infants born with malformations. Though the clusters of severe birth defects resulting from this outbreak have subsided, ZIKV continues to be a concern throughout much of Latin America and the Caribbean. Travel and sexual intercourse remain the dominant transmission risk factors for women of reproductive age and their partners. This is particularly true for communities in Brooklyn, New York, that comprise large immigrant and foreign-born populations. Practitioners of public health understand little about how women at risk for ZIKV are most likely to receive information about the virus or who they trust most to provide that information. In the context of five focus group discussions, this study explored the knowledge and communication preferences of 20 women of reproductive age in Central Brooklyn. Results derived from a thematic analysis suggest that while most women are familiar with mosquitos as ZIKV vectors, knowledge of sexual transmission is considerably lower. Many respondents believe that only women who are pregnant or trying to become pregnant are at risk, and public health agencies, such as the U.S. Centers for Disease Control and Prevention, remain the most trusted sources of information. These findings can support more effective communication about the risks of ZIKV infection and other vector-borne diseases to women in New York City and similar urban communities.

## Introduction

Zika virus (ZIKV) is the causative agent of a vector-borne disease that is spread primarily by the bite of infected *Aedes aegypti* and *Ae. albopictus* mosquitos and is also sexually transmitted.^1^ Most symptoms are generally mild and include fever, rash, muscle and joint pain, and headaches. Complications from ZIKV infection include microcephaly, a severe birth defect involving brain damage in the offspring of infected women, and Guillain-Barré Syndrome, an autoimmune disorder that can lead to peripheral nerve damage and ultimately partial paralysis.^2^ The disease gained international attention in 2015 when the virus infected millions throughout Latin America and the Caribbean, prompting the World Health Organization to declare the outbreak a Public Health Emergency of International Concern in February 2016.^3^ More than 50 countries reported ZIKV cases in a matter of months, and many local epidemics were extraordinarily pervasive. The rapid spread was likely due to a combination of factors including a large susceptible population, a hospitable climate for the mosquito vector, and sexual transmission.^4^

ZIKV became a nationally notifiable condition in the United States in 2016.^5^ Health campaigns and education have been ongoing in the United States ever since.^7^ While early interventions included expanded mosquito vector larviciding and adulticiding programs throughout many high-risk communities in the US, most interventions after 2017 have relied solely on risk mitigation via targeted health communication campaigns.^8,9^ The CDC notes several important goals of Zika communication activities including increasing overall awareness of the virus, educating pregnant women, and educating women of reproductive age.^10^

Numerous studies, however, have demonstrated a clear gap between what was communicated by practitioners of public health and what is known by the public when it comes to engagement in personal protective measures.^11,12^ This is especially true for ZIKV. In one study, researchers found that the vast majority of participants (97%) did not have knowledge of all routes of transmission and nearly three in four people (73%) did know that condom use was a key prevention measure.^13^ Other studies have also identified gaps related to vector habits and environmental controls.^14–16^ Messages need to be timely, culturally appropriate, and easy to understand for risk communications to be effective. To more effectively communicate what is known to the public, we must improve our communication efforts and prioritize potentially vulnerable populations who are at greater risk of exposure yet can be less likely to engage in preventive practices due to gaps in knowledge.^17^ We will not be able to effectively address these issues until we understand how to appropriately tailor communications and public health interventions.

The purpose of this qualitative study is to explore the preferred methods, modes, and frequency of communicating about ZIKV among women of reproductive age in central Brooklyn. To guide this research, we posed two research questions: 1) How do women of reproductive age in Central Brooklyn receive information about Zika virus? and 2) How do women of reproductive age in Central Brooklyn prefer to receive information about Zika virus going forward? The study explored several domains. Preferred methods, frequency, and modes of communication to address gaps in ZIKV knowledge were all explored. We aimed to gain meaningful insights on group consensus via participant focus group discussions (FGDs) and in-depth contextual analysis. We conducted this research to explore perceptions and preferences and solicit rich information on ZIKV from the perspective of a group of study participants who are most vulnerable to the disease. We used focus group discussions because we wanted to provide a platform for an exchange of viewpoints and ideas, and assess where agreements and disagreements occurred. Our thematic analysis was guided by the principles in the WHO Strategic Communications Framework including accessibility, trustworthiness, and relevance.

The study was among women of reproductive age who reside in Central Brooklyn, in New York City. Central Brooklyn consists of several neighborhoods comprising different ethnic groups. Though the community is situated in one of the most affluent cities in the world, residents of Central Brooklyn tend to be poorer and sicker than their counterparts across New York and the United States in general.^19,20^ This is largely due to various social determinants of health (e.g. income, education, occupation, housing, food stability), and potential travel patterns.^21–23^ More than 75% of the community identifies as Black including West Indian and Afro-Caribbean or Latinx, and approximately 42% – higher than the Brooklyn average of 37.5% – is foreign-born.19,20 Women of reproductive age can be particularly vulnerable to the effects of ZIKV, given the risk of birth defects and potential for sexual transmission.^24^ It is therefore beneficial to learn what this population knows about ZIKV and how they would like to receive information to fill their knowledge gaps. Assessing current and preferred methods of receiving knowledge can cost-effectively inform interventions to reduce the likelihood of future infections and improve population health.

### Theoretical Framework

The World Health Organization’s Strategic Communications Framework for effective communications guided this research.^18^ The framework posits that there are six guiding principles for effective communications: accessible, actionable, credible and trusted, relevant, timely, and understandable. We focused on four principles from the framework to guide this research: accessibility, credibility and trustworthiness, relevance and timeliness. We aimed to identify the most widely utilized channels and modalities for receiving information about ZIKV to approximate accessibility and the most credible and trusted sources for providing information. We also assessed the relevance and timeliness of messages both during the previous ZIKV epidemic and going forward by asking participants not only what they wanted to learn more about but how often they would like to receive that information. Furthermore, additional information on knowledge and perceived vulnerability were also collected. The research aimed to provide a basis for tailoring messages based on this population’s current levels of knowledge to protect a potentially vulnerable population. Recommendations made are based on thematic analysis of qualitative data collected directly from the study population to ensure they are in a preferred format and are easily understandable.

## Methods

### Participant Selection and Recruitment

We purposively sampled respondents by considering age and race in an attempt to represent various opinions and attitudes about ZIKV. Participants were eligible to participate if they met the following inclusion criteria: 1) a woman between 18-45; 2) a current resident of Central Brooklyn; 3) had heard of ZIKV; 4) willing to review an information sheet about the research; 5) had stable internet connection at home; 6) had access to a laptop or phone with a camera; and 7) had access to a private space to conduct the interview. A potential participant was ineligible to participate if they did not meet all inclusion criteria.

Facebook’s paid advertising feature was used to reach respondents from the study population. Advertisements were targeted to women 18-45 who reside in Central Brooklyn. The advertisements mentioned the purpose of the research and a cash incentive participated in a focus group discussion. The advertisements also contained a link to a screening questionnaire housed in REDCap that collected self-reported information confirming each respondent met the aforementioned inclusion criteria. Focus group discussion participants were initially contacted by email after expressing interest through the REDCap screening questionnaire.

### Data Collection

Once the participant confirmed interest, we gathered additional information on race/ethnicity, age, and place of birth. Potential participants were offered three timeslots to choose from and were asked to select their top two. We aimed to have an average of four respondents per discussion – a recommended amount for Computer Mediated Communication (CMC)-led focus group disucssions.^25^ We scheduled five participants per timeslot, assuming one might not be able to make it at the last minute. A final reminder was sent out on the morning of each discussion that contained a link to a Doxy.me account.^26^ If a respondent missed their initial timeslot, they were given the option to sign up one more time only. Each discussion lasted approximately one hour and a thank you email was sent within one day of the discussion that contained confirmation of a $30 incentive payment and a link to more information about ZIKV from the US Centers for Disease Control and Prevention. Audio recordings of each session were obtained and stored in a password-protected cloud-based server.

### Study Design

A semi-structured focus group discussion (FGD) guide was informed by the research questions and reviewed in consultation with qualitative research and infectious disease experts during the piloting phase. Topics explored in the FGDs were: 1) what respondents have heard about ZIKV including knowledge of transmission and prevention; 2) how respondents have received information about ZIKV in the past; 3) who they trust most to provide information about ZIKV; 4) how and how often they would like to receive information about ZIKV transmission and its risks going forward. Sessions were moderated by one of the authors of this study and we obtained audio recordings of each FGD. Participants received a $30 incentive for their participation.

### Data Management and Analysis

Once all focus groups were completed, we utilized a transcription service to convert audio recordings to text verbatim including pauses, laughter, and nonverbal cues when available (e.g., nodding when verbally acknowledged by the facilitator). The final transcribed documents were separated into twenty unique documents, one corresponding to each participant. This allowed us to attach demographic characteristics to each participant in Dedoose v8.0.42 (https://www.dedoose.com) for qualitative analysis—a feature that is otherwise unavailable for group transcripts.^27^ This also allowed us to compare across individuals rather than across focus groups, which were assigned at random.

A thematic analysis was then conducted to assess patterns of meaning in the participants’ receipt of knowledge in addition to their communication and intervention preferences. Initial root codes were created based on each of the semi-structured open-ended interview questions posed during the FGDs. Emerging codes (subcodes in Dedoose), on the other hand, were largely created in the analysis phase of the study directly from the data.

Data were interpreted via thematic analysis guided by important concepts for analysis found in WHO’s Strategic Communications Framework for effective communications. Interview questions were informed by the research questions and connected to the four principles of interest from the strategic communications framework: accessibility, credibility and trustworthiness, relevance, and timeliness. Codes and root codes were then established using the interview questions. Codes were counted using Dedoose and weighted according to the importance of the response; responses that were categorized by the coder as ‘particularly important, interesting or relevant’ were given an additional point to ensure the findings were not biased toward the thoughts of more talkative respondents. Those codes with the greatest weighted counts were then considered for inclusion in the thematic analysis, and appropriate quotes were identified to support each theme.

### Ethics Statement

This study was reviewed by the SUNY Downstate’s Office of Research Administration Institutional Review Board (IRB) and received an exemption.

## Results

In total, 59 respondents expressed interest in participating in a focus group discussion. All 59 were screened and 30 that met the inclusion criteria were contacted. Focus group discussion participants were recruited from December 2020 to January 2021. A sample size of 20 was finalized when data saturation was reached. Five focus groups with an average of four participants (range: 3-5) each were held between December 2020 and January 2021 (Table 1).

Sixty codes were created and applied 634 times to 207 unique excerpts. Based on the patterns identified in the analysis, six concepts were analyzed: 1) knowledge of ZIKV; 2) past receipt of information; 3) perceived susceptibility or vulnerability; 4) trusted sources of information; 5) preferred medium/modes of communication for receiving information going forward; and 6) preferred frequency for receiving information going forward. The themes that emerged from these concepts are discussed below.

### ZIKV Knowledge

We explored knowledge of ZIKV by asking about transmission, prevention, and population vulnerability. Knowledge as an important theme often included information on where the virus first emerged, where it is currently circulating, and the 2016-2017 epidemic in NYC. Several participants also discussed information on vaccines and therapeutics.

> *“Women that were pregnant were… giving birth to children and I think that I recall the most, they were mentioning that women that were infected with Zika virus were giving birth to children that had oddly shaped heads*.*”*
>
> *“I remember that it really affected pregnant women, and, if you were infected, there was a high chance that you could miscarry, or your child might be born with deformities*.*”*
>
> *“I remember that it spread by mosquito bites and how they were spraying in Brooklyn for it*.*”*

When it comes to knowledge of ZIKV, the respondents most often agree that the virus affects pregnant women, is associated with travel, and is transmitted by mosquitos. Respondents demonstrated knowledge on a range of topics including transmission, environmental controls, personal protective measures, health outcomes, and vulnerable populations. Most respondents talked about microcephaly and birth defects in general. Knowledge of sexual transmission in this series of focus groups, not mentioned in the above excerpts, appeared less common than knowledge of vector-borne and vertical transmission, as it was mentioned many fewer times and numerous participants reported that they had never heard of this mode of transmission. The simplest theme emerging from the findings was that knowledge of adverse health outcomes was commonplace among participants. Though arguably less important than knowledge of transmission or personal protective measures, discussions of associated birth defects dominated most of the conversations. This implies that knowledge, which was largely tied to memories of the last Zika epidemic, was linked to the more graphic outcomes of ZIKV often focused on by the media.

### Sources of ZIKV Information

We asked how participants have received information about ZIKV in the past by probing on two concepts: from whom they received the information and how the knowledge was transferred. Most responses for the “who” fell into the following categories: CDC or Department of Health, a doctor, nurse, or other healthcare professional, a family member or friend, professors, or the media. The theme of information receipt was often directly linked to the perceived credibility of the source of information. The “how” was often through television or online advertisements, social media, print media (e.g. newspapers), or physical advertisements (e.g. pamphlets, posters).

> “I [remember hearing about it in] the New York Post and Huffington Post and USA Today…and CNN. So those are the outlets that I use on a daily basis.”
>
> “A few years ago, I was having my third child, and I remember seeing in the midwife office that I go to, there would always be posters about Zika virus and traveling.”
>
> “When you’re going to the airport to fly, you would see a poster all the time like just telling you to beware, coming to and from the airport.”
>
> “I got information from a reproductive endocrinologist, or a fertility specialist, because that who I was seeing at the time.”

Most respondents reported that they have received information about ZIKV from the news, physical advertisements, and/or healthcare professionals (including friends who work in healthcare). Accessibility and media preferences largely dictated receipt of information, as did personal circumstances at the height of the epidemic. Participants who were pregnant at that time largely sought out or received information from personal OB/GYN’s or fertility specialists, whereas frequent travelers would often turn to official government sanctioned websites for information. The type of information sought out was often closely linked to the personal risk profile of the recipient.

### Perceived ZIKV Infection Susceptibility

Perceived susceptibility to ZIKV was explored by asking who the participants thought was at the greatest risk of infection or severe health outcomes. The prompt was intentionally left vague to leave room for interpretation. Participants could have provided responses that looked at differences by gender, race, ethnicity, socioeconomic status, geography and more.

> “For me, I always perceived the information as ‘if you are not a pregnant woman, you do not need to worry about this virus’”.
>
> “It didn’t fit my demographics, because I wasn’t a pregnant woman. So, I didn’t really pay that much attention to it.”
>
> “I was thinking of getting pregnant [at the time], and I was looking to travel with my husband. So, we had a long conversation about the Zika virus and countries I should be thinking of traveling to.”
>
> “I feel like women were more concerned than men.”
>
> “We weren’t at [childbearing] age. And even if we were at the age, if you’re not pregnant, or you’re not thinking about being pregnant, you don’t feel concerned because it doesn’t have a direct impact on [you].”

The general consensus among the participants was that you did not need to worry about ZIKV unless you were a woman who was pregnant or looking to become pregnant. Several participants also mentioned risk by geography, specifically related to Latin America, the Caribbean, and Africa. No participants discussed risks by race, ethnicity, or socioeconomic status. This suggests the media’s focus on microcephaly might have inadvertently altered perceived susceptibility to the virus. Though severe, birth defects are considered a rare outcome of ZIKV. Even so, many participants focused on these outcomes and their connection to new mothers when discussing susceptibility. More common outcomes like headaches, joint pain, and rashes were often overlooked.

### Trusted Sources of Information

Trust is paramount in risk communications, especially when working with vulnerable populations. The concept of ‘trust’ was gauged by asking participants who they trusted most to provide information about ZIKV. Responses included groups of people such as medical personnel, specific individuals like personal physicians, or health agencies like the US CDC.

> “I definitely [trust] CDC. Although I wasn’t looking for guidance on Zika. [But] I do now for COVID. So, I definitely—CDC’s my bible.”
>
> “There’s national information given by the CDC, but I wanna know a little bit more. Because New York City is definitely very unique, so I would say I trust DOHMH, a great amount as well as New York City Health and Hospitals.”
>
> “I personally trust only either medical professionals or you know websites or sources coming directly from a reputable medical background.”
>
> “I trust my doctor only.”

When asked who they trusted the most to provide information about ZIKV, participants most often mentioned public health agencies including the United States Centers for Disease Control and Prevention (US CDC) or a local department of health. Many of the above excerpts and several others highlight the importance of federal and local government in providing clear and credible risk communications during an epidemic. This was followed closely by doctors, nurses, or other healthcare personnel. The common theme among trusted sources of information was expertise. Participants often relied on media sources that referred to an array of experts including public health, medical, and laboratory scientists.

### Receiving Information

Messages should not only be tailored to a study population, but also channeled through an appropriate mode of communication that will be both widely available and accepted by the study population. Receipt of preferred modalities for information on ZIKV in the going forward was therefore explored to understand the channels through which the participants would like to receive specifically unsolicited information now and in the future. We made no mention of the rates of infection or potential messenger. Instead, we allowed participants to widely interpret the prompt and provide feedback on whichever modalities they think would be most effective to reaching them personally.

> “I think having a text is the most in my face kind of thing, because everything else is so easy to ignore.”
>
> “I think getting up to date information either via text message or through an app [would be best]. I have the Citizen App, and I certainly use that when I’m looking at what’s going on in my neighborhood. It doesn’t require me to seek anything out or have to remember to look for the information, it just, you know comes to me. And I think that’s ideal… as long as it’s coming from a reputable source that I can trust.”
>
> “I know we’re very paperless, and I may be traditional this way, but I think I look more at things that come in the mail, like than email.”
>
> “[Having] ads come on Twitter…and medical Instagram pages, or Facebook pages is best. Because a lot of people, especially like in our generation, are always on social media.”

There was no one consensus about how the respondents would like to receive information about ZIKV going forward. Text, email, printed materials, online or television advertisements, and the news were all frequently mentioned potential modalities. Participants were fairly evenly split between typical modes of health communication such as printed materials sent from doctors’ offices or health departments and more technology-driven materials such as messages from health-based apps. Information receipt will have to be varied and include a diverse array of modalities.

### Frequency of Information

Finally, preferred frequency of unsolicited communications about ZIKV was explored by asking how much participants would like to hear about the virus going forward. Regular (e.g. bi-monthly, quarterly) updates about an emerging virus can be useful to help update the public on important information and synthesize pertinent lessons learned.

> “[Not] unless there’s something that I personally need to like follow up on. Like if there’s, additional precautionary things that I should be doing.”
>
> “[Not] unless there’s been a change in something. Like if it’s in a year from now and… it’s on the rise. Other than that, we’re just gonna get blindness to it.”
>
> “I feel like if there’s a drastic spike, like there’s an increased number of cases. Or even when there’s a dramatic decrease, I feel like that’s the time when you should alert individuals.”
>
> “I would say I would wanna hear about it if there was a significant change. So if there’s a drastic spike, or if it’s the opposite. If there’s been some sort of change for the positive, that it’s no longer a concern, or there’s been some sort of medical discovery that’s helped with preventing it. Anything like that I don’t think I would think it’s information overload.”

There was a consensus on the frequency of future communications about Zika, with participants overwhelmingly preferring to only receive information where there were notable updates about Zika, good or bad (e.g., an increase in the prevalence of Zika, new research findings, a lifting of travel warnings). While numerous studies have assessed current methods and modes of receiving information and most trusted sources of information, few, if any, have explicitly asked respondents how frequently they would prefer to receive information about ZIKV going forward. There was a general sense that participants were subject to information overload given recent epidemics and public health emergencies of international concern including COVID-19, Ebola, MERS, H1N1, SARS and more. Though regular updates on diseases like ZIKV can empower recipients of the information and improve population knowledge, communications should be infrequent and focused on transfer of new and notable information only.

## Discussion

In this study, we found that most participants would like to receive information about ZIKV via text, email, and printed materials only when there is some sort of notable update about the virus. Healthcare staff and leading health agencies such as the US Centers for Disease Control and Prevention are the most trusted providers of information, and participants believe that women who are pregnant or trying to become pregnant are not only at greatest risk from ZIKV but often the only group at risk. Knowledge of sexual transmission was considerably lower than that of mosquito transmission. Generally, the FGD participants presented considerable knowledge of mosquito and vertical transmission of Zika. This appears largely consistent with findings from other studies assessing knowledge, attitudes, and practices in or around New York City.^13,28,29^ Knowledge of sexual transmission was less common, though results from published research vary.

When it comes to past receipt of information, the news and healthcare professionals were preferred sources of information, though important differences were uncovered by age and behaviors. While there have only been a few studies that published data on how people receive information about ZIKV, this research appears to echo the findings from a seminal study published by Piltch-Loeb and Abramson in 2017.^33^ One notable difference is that current respondents appeared less willing to receive information about ZIKV from family, friends, or social media unless they were in some way connected to a medical pursuit. Our findings are also consistent with the findings of a community-based participatory research study by Juarbe-Rey et al. in 2019, which found that nearly three in four respondents had received information about ZIKV through television including the news.^34^ In general, younger respondents (e.g. those in their twenties) were more likely to have received information through the internet—especially social media sites including Facebook and Twitter. This is consistent with findings from Guo et al., which found that 71.1% of respondents in a sample of 492 pregnant women first heard about the virus from the internet.^35^ The median age in that sample was 30 years. These findings were also echoed in another article by Ramisetty-Mikler and Boyce, which found that 65% of respondents used the internet as the main source of information for ZIKV.17 The issue of misinformation on social media did arise in at least two of the FGDs. In general, younger participants appeared more confident in their abilities to detect and navigate around sources of misinformation when compared to older participants.

The majority of studies on ZIKV were published between 2017 and 2019, immediately following the declaration from the World Health Organization that ZIKV was a ‘Public Health Emergency of International Concern’. Since that time, concern for the virus has fallen precipitously in nearly all groups studied. For example, our findings are consistent with those of Ellingson, Bonk, and Chamberlain (2017), which found reduced concern and fewer self-set travel restrictions over time.^36^ Numerous studies have also found that pregnant women are more knowledgeable and likely to engage in preventive practices when compared to non-pregnant women, which has a similar conclusion to the findings outlined above.^13^ Furthermore, the study by Plaster et al. (2018) confirmed these findings, with 73.1% of respondents agreeing that ‘Zika virus is a serious health condition for women’, and only 30% of respondents agreeing that ‘Zika virus is a serious health condition for men’.30 Given the increase in news coverage surrounding the large clusters of children with microcephaly born between 2016 and 2017, it is unsurprising that many FGD participants saw pregnant women and their unborn children as most vulnerable to infection or adverse health outcomes from Zika. The participants generally agreed that men were not considered vulnerable populations, though this is likely due to the fact that many were unaware that ZIKV could be sexually transmitted. Discussions of race or socioeconomic status as they pertain to vulnerabilities did not arise in any of the focus groups.

When it comes to considering which sources of information they trust most, most FGD participants mentioned healthcare professionals and federal or local health agencies such as the US CDC or the New York City Department of Health and Mental Hygiene. While it is encouraging to see how many people in these focus groups trust the US CDC and the New York City Department of Health and Mental Hygiene (DOHMH), this is in direct contrast to findings from a study by Berenson et al. (2017), which found that only 20% of U.S. born respondents and 9.6% of respondents born in countries with ongoing ZIKV transmission trusted government announcements.^22^ This also differs considerably from another study by Poorasingh et al.^37^ That study, which interviewed 91 respondents in antenatal care clinics in Trinidad and Tobago, found that fewer than one in four (23.1%) listed health agencies such as the Pan American Health Organization as a top trusted source of information. It is possible that, given the rise in COVID-19 throughout much of 2020 and the general public’s reliance on federal and local health authorities, my findings are biased. Furthermore, the rise of misinformation over the past five years could be driving this move toward more official sources of information. Fortunately, we did identify one study that validated our findings, with 73% of respondents turning to the CDC’s website as their trusted source of information.^38^

Several studies have assessed preferred modalities for receiving information about ZIKV.^17,36,38^ Like many of these studies, our research did not identify one specific modality that was favored by all participants. Rather, several preferred modes of communication were discussed consistently throughout the focus group discussions, including texts, emails, and printed materials like educational brochures or flyers. This highlights the importance of utilizing a diverse approach to communication modalities. One study by Ellingson, Bonk, and Chamberlain (2018), found that educational brochures were the preferred source of information.^38^ Emails and websites were also considered preferred sources, though this study did not explicitly ask about health-based apps. Medical Facebook or Twitter pages were perceived as undesirable sources of information, which differs slightly from the findings in our research. There appeared to be the greatest interest in physical advertisements such as subway posters or health center pamphlets that were specifically developed or endorsed by trusted medical or healthcare groups like a family practice or the New York City Department of Health and Mental Hygiene. Many FGD participants agreed that auto-generated texts or emails would likely be missed due to information overload, particularly given the COVID-19 pandemic. Though initially surprised by the desire to receive educational brochures when most people have the capability to receive information digitally, our findings are consistent with findings from Ellingson, Bonk, and Chamberlain.^38^

This article had several limitations. Our FGD participants could be slightly demographically different than the residents of Central Brooklyn. While we did not explicitly ask about ethnicity, education, or socioeconomic status, white participants represented a disproportionately large share of the racial breakdown. Furthermore, participants were recruited solely via Facebook advertisements, omitting the perspectives of those without an account and leading to documented selection bias.^39^ Participants also needed to have a stable internet connection and a private space to be included in a focus group, potentially limiting comparability with Central Brooklyn residents who are lower income. Finally, FGDs were held only in English, though many Brooklyn residents speak Spanish, French, Haitian Creole, and other languages.

This study also has several strengths. We received more interest in the research than was expected, which allowed for a purposeful selection of participants to ensure we had a wide age range represented. Race and age, in particular, were considered when selecting participants. Conducting the focus groups remotely resulted in high (20/21) participation rates among scheduled participants. To our knowledge, this is the first study that assessed unsolicited communication frequency preferences related to ZIKV in the published literature. Furthermore, the results can apply not only to similar populations but also to similar vector-borne diseases such as Dengue. Finally, while studies utilizing CMC as a platform to hold focus group discussions have encountered participants who have technical challenges accessing the discussion, there is often broad agreement that CMC allows for greater anonymity and ease of scheduling when compared to in-person discussions.^40^ Furthermore, comparisons of in-person versus CMC focus groups have found that perceived anonymity across the internet can stimulate group discussion and disclosure, often allowing for even greater sharing of ideas during CMC.^41^

### New Contribution to the Literature

We were unable to identify any study that specifically explored the preferred frequency for receiving information about ZIKV going forward. However, these findings are likely consistent with the population at large for two reasons. First, numerous studies have demonstrated decreasing concern in ZIKV over the past few years. This would also certainly translate to a reduced desire to receive information about ZIKV. Finally, participants in this study were nearly unanimous in their desire to receive information only when there was a notable change in Zika, such as an increase in cases.

We recommend that future public health communications about ZIKV go beyond these topics to include important and often overlooked modes of transmission like sexual contact, even among partners of women of reproductive age who have traveled to Zika-endemic regions. Messages should also focus on preparedness and engagement in personal protective measures rather than threat or fear-based messaging by trusted messengers such as medical professionals or local health agencies, as they have been demonstrated to be more effective in producing increased levels of public engagement.^43,44^ Finally, it is important to ensure that we are not only communicating through the appropriate channels and sources but also highlighting the correct messages. It was noted in 2017 that the New York City subway system was filled with posters showing big mosquitoes, but nearly all cases in New York were travel-associated or sexually transmitted.^45^

## Conclusions

While most respondents are familiar with mosquito transmission of Zika, fewer were aware of sexual transmission, and many respondents believed that only women who are pregnant or trying to become pregnant are at risk. Additionally, medical professionals and public health agencies remain the most trusted sources of information, and communications should be tailored through several modalities such as texts, emails, and brochures to account for the wide range of communication preferences. Public health practitioners can utilize this information to effectively communicate about the risks of ZIKV and other vector-borne diseases to women in Central Brooklyn and similar urban immigrant communities by channeling messages through trusted messengers and widely accepted modalities. These findings can guide public health practitioners to more effectively communicate with existing vulnerable populations who either travel or have spouses who travel to areas with ongoing ZIKV transmission. Additionally, it can provide important insights into this population or comparable at-risk urban populations for future ZIKV epidemics or similar vector-borne diseases such as Dengue. Further studies are needed to design and test future messages and educational materials to ensure that they are culturally appropriate and impactful.

## Data Availability

All data produced in the present study are available upon reasonable request to the authors

## Acknowledgments

We thank Tracey Wilson, PhD for her guidance during the earlier stages of this work.

## Notes

**Funding Statement** This work was supported by the Lenard and Christine Szarek Fellowship conferred on Russell Dowling.

**Conflict of Interest** The authors have no conflicts of interest to declare.

### Competing Interest Statement

The authors have declared no competing interest.

### Funding Statement

This study was funded by the Lenard and Christine Szarek Fellowship conferred on R.A.D.

### Author Declarations

IRB of SUNY Downstate Health Sciences University waived ethical approval for this work

